# DEMOGRAPHIC & CLINICAL PROFILE OF PATIENTS PRESENTED WITH ACUTE MYOCARDIAL INFARCTION DURING COVID 19 PANDEMIC - A TERTIARY CARE CENTRE STUDY

**DOI:** 10.64898/2025.12.09.25341947

**Authors:** Deepa Noronha, H Haroon, Padmanabh Kamath, D Narasimha Pai, Chaithra Nayak, Mishika Chawla, Aastha Pahuja, Reema Dsouza

**Author notes:** **Corresponding author 1:** Email id. Email id.

## Abstract

**Background:** Coronavirus disease 2019 (COVID-19) has had a profound impact on cardiovascular health, influencing both the presentation and outcomes of acute myocardial infarction. This study aimed to evaluate the demographic and clinical profiles of patients with ST-segment elevation myocardial infarction (STEMI) presenting to a tertiary care center during the COVID-19 pandemic and to compare early and late presenters in terms of outcomes.

**Methods:** A retrospective cohort study was conducted on 885 patients diagnosed with STEMI during the COVID-19 pandemic. Demographic, clinical, electrocardiographic, echocardiographic, and coronary angiographic data were collected. Patients were classified as early presenters (<12 hours) or late presenters (>12 hours) on the basis of the time from symptom onset to hospital arrival. Statistical analyses were performed via independent t tests and chi-square tests, with p < 0.05 considered significant. This study aligns with United Nations Sustainable Development Goal (SDG) 3: “Good Health and Well-Being”.

**Results:** The mean age was 58.45 ± 10.61 years, with males comprising 76% of the cohort. Diabetes mellitus (75%) and hypertension (53%) were the most prevalent risk factors. Among the ECG findings, anterior wall MI (57%) was most common. Single-vessel disease was predominant among early presenters (67%), whereas double/triple-vessel disease was more common among late presenters (46%). In-hospital mortality was 4.18%, which was significantly higher in late presenters (6%) than in early presenters (3%). Non survivors were older (65.2 ± 12.8 years) and had higher rates of hypertension and heart failure symptoms (p < 0.05).

**Conclusion:** During the COVID-19 pandemic, delayed hospital presentation among STEMI patients was associated with more extensive coronary disease, higher complication rates, and increased mortality. Early recognition and timely intervention remain crucial for improving outcomes in acute myocardial infarction patients during healthcare crises.

## INTRODUCTION

Coronavirus disease 2019 (COVID-19) represents the clinical presentation caused by infection with severe acute respiratory syndrome coronavirus 2 (SARS-CoV-2). Transmission occurs predominantly via respiratory droplets expelled when infected individuals cough or sneeze, most frequently from symptomatic cases, although increasing evidence indicates that individuals without symptoms can also contribute to spread. Clinically, the disease often presents with significant respiratory symptoms—fever, cough, and fatigue—and may advance to severe complications such as pneumonia, acute respiratory distress syndrome (ARDS), and circulatory shock. Beyond pulmonary involvement, COVID-19 has been associated with notable cardiac complications.

Several mechanisms have been proposed to explain how the pandemic has influenced cardiovascular health trends. Both pre-existing cardiovascular disease and traditional cardiovascular risk factors are linked to poorer outcomes in infected patients. SARS-CoV-2 may disrupt endothelial integrity and impair cardiomyocyte function, and reports describe conditions such as myocarditis, thromboembolic events, and widespread vascular inflammation. Additionally, indirect consequences of the pandemic—such as reduced access to routine medical care—may further contribute to adverse cardiovascular outcomes. The severity of cardiac injury appears to be influenced by viral load, the intensity of the host immune response, and the presence of underlying health conditions.

Research by Liu PP et al. (2020) highlighted that persistent immune activation in susceptible individuals, particularly older adults and those with cardiovascular risk, may trigger a hemophagocytic-like inflammatory state, characterized by uncontrolled cytokine release that can culminate in multi-organ failure and death. Early involvement of the cardiovascular system is often indicated by elevated levels of high-sensitivity troponin and natriuretic peptides, which, along with inflammatory markers such as interleukin-6, have been shown to predict poor clinical outcomes when their concentrations continue to rise during infection.

The purpose of this study was to evaluate the demographic and clinical data of patients with ST segment elevation myocardial infarction presenting to tertiary care centers during the COVID-19 pandemic. Our study evaluated various electrocardiographic patterns in patients with ST segment elevation myocardial infarction and assessed echocardiographic and coronary angiographic findings. This study aligns with United Nations Sustainable Development Goal (SDG) 3: “Good Health and Well-Being”.

## METHODS

Study design and study area: This retrospective cohort study was conducted tertiary care hospital in the coastal Karnataka region after approval was obtained from the Ethics Committee of our institute. A total of 885 individuals who were diagnosed with ST segment elevation myocardial infarction during the COVID-19 pandemic were included in this study. The data were accessed through medical records and a system-based search that included demographic, clinical and diagnostic test results.

Data collection methodology: Demographic data such as patient age, sex, height, weight and BMI were collected from medical records. During the COVID-19 pandemic, all patients hospitalized with COVID-19 were given COVID-19 tests as part of the protocol. For our study, patients were categorized based on their time of presentation, i.e., early presenters (<12 hours) or late presenters (>12 hours). Risk factors such as diabetes mellitus, hypertension, a history of smoking, dyslipidemia and a family history of ischemic heart disease were assessed. Baseline 12-lead electrocardiographic, 2D echocardiographic and coronary angiographic findings were documented. Clinical findings, including heart rate, blood pressure, and baseline oxygen saturation, and laboratory findings, such as the status of cardiac biomarkers and serum electrolytes, were also collected. From these findings, the TIMI risk score for ST elevation myocardial infarction (STEMI) was calculated. The interval from the onset of chest pain to first medical contact was documented. The treatment strategy for acute myocardial infarction patients was documented. Data regarding in-hospital cardiac and noncardiac complications were assessed through medical records and documented.

Statistical analysis: An independent sample t test was used to analyze continuous data, whereas a chi-square test was used to analyze categorical data. The means and standard deviations were used for numerical data, whereas percentages and numbers were used for categorical data. A p value of <0.05 was considered to indicate statistical significance. Jamovi version 2.6 was used to analyze the data.

## RESULTS

This study included 885 patients who were diagnosed with ST segment elevation myocardial infarction during the COVID-19 pandemic in a tertiary care center. The mean age of the study population was 58.45 ± 10.61 years, ranging from 27--85 years of age, 76% of whom were males. The mean body mass index (kg/m^2^) was 23.88 ± 2.62, and the mean body surface area (m^2^) was 1.52 ± 0.105. Among the risk factors, 660 (75%) patients had diabetes mellitus, and 470 (53%) of the population were hypertensive, which was more common in the male population 505 (76%) and 344 (73%) patients, respectively). All other risk factors are also predominant in the male population. The demographic details and various risk factors are depicted in Table 1.

**Table 1:**
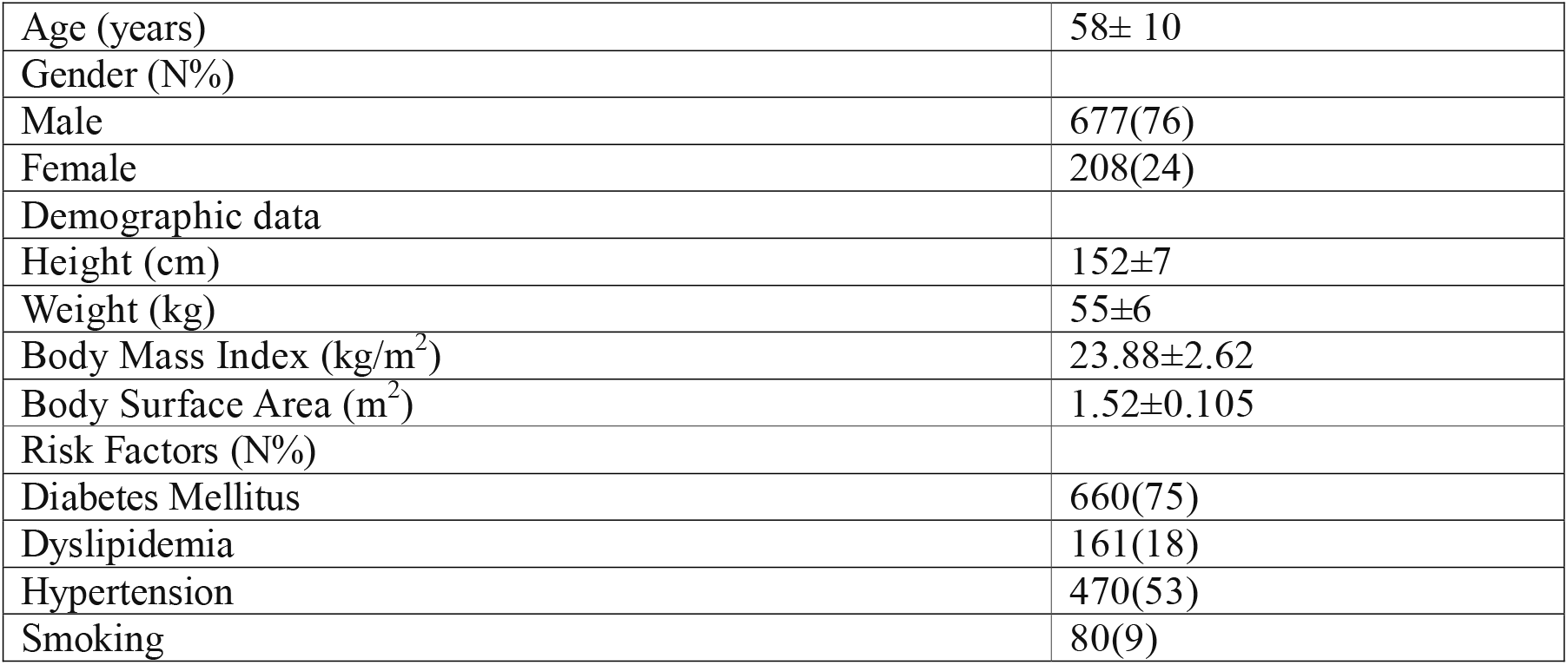
Demographic profile of the patients.

Among the study population, QRBBB was detected in 148 (17%) patients, complete heart block was detected in 119 (13%) patients, LBBB was detected in 104 (12%) patients, and sinus bradycardia was detected in 95 (11%) patients via ECG. Among the 885 patients, 504 (57%) had ST segment elevation in the anterior leads and were diagnosed with AWMI, 339 (38%) had inferior lead changes in the IWMI, and 42 (5%) had lateral lead changes and were diagnosed with inferolateral wall MI. Among the study population, 858 patients were subjected to coronary angiography. Among the 532 patients (62%) with single-vessel disease, 243 (28%) had double-vessel disease, and the remaining patients had triple-vessel disease. The ECG findings are depicted in Table 2.

**Table 2:** ECG findings and coronary lesion types.

| ECG Findings | Early Presenters<br>(n=510) | Late Presenters<br>(n=375) | p value |
| --- | --- | --- | --- |
| CHB | 75(15%) | 44(12%) | 0.231 |
| LBBB | 53(10%) | 51(14%) | 0.169 |
| QRBBB | 77(15%) | 71(19%) | 0.145 |
| Sinus Bradycardia | 49(15%) | 46(12%) | 0.227 |
| ECG Diagnosis |  |  |  |
| AWMI | 275(54%) | 229(61%) | 0.043 |
| IWMI | 205(40%) | 134(36%) | 0.043 |
| Infero lateral Mi | 30(6%) | 12(3%) | 0.043 |
| **Coronary Lesion Type** |  |  |  |
| SVD | 336(67%) | 196(55%) | <0.05 |
| DVD | 129(26%) | 114(32%) | <0.05 |
| TVD | 34(7%) | 49(14%) | <0.05 |

We analyzed in-hospital complications and their outcomes during the hospital stay. Among the 44 patients who had complications, 29 (4%) had cardiac arrest, and 4 had ventricular tachycardia and cardiogenic shock. We also noted in-hospital deaths in 37 patients. The patient outcomes are depicted in Table 3.

**Table 3:** In-hospital complications.

| **In-Hospital Complications** | **Early Presenters (n=510)** | **Late Presenters (n=375)** | **p value** |
| --- | --- | --- | --- |
| Asystole | 1(0.2%) | 0 | 0.086 |
| Cardiac arrest | 10(2%) | 19(5%) | 0.086 |
| Cardiogenic shock | 3(0.6%) | 1(0.3%) | 0.086 |
| Cardio-pulmonary arrest | 1(0.2%) | 0 | 0.086 |
| Free wall rupture | 0 | 1(0.3%) | 0.086 |
| Pericardial effusion | 0 | 1(0.3%) | 0.086 |
| Pulmonary edema | 1(0.2%) | 1(0.3%) | 0.086 |
| Ventricular septal rupture | 1(0.2%) | 0 | 0.086 |
| Ventricular tachycardia | 2(0.4%) | 2(0.5%) | 0.086 |

Based on the clinical outcomes of patients observed during their hospital stay, the entire population was grouped into survivors and nonsurvivors. Among the 885 patients, 848 (96%) survived, and the remaining patients died in the hospital. The details are depicted in Table 4.

**Table 4:**
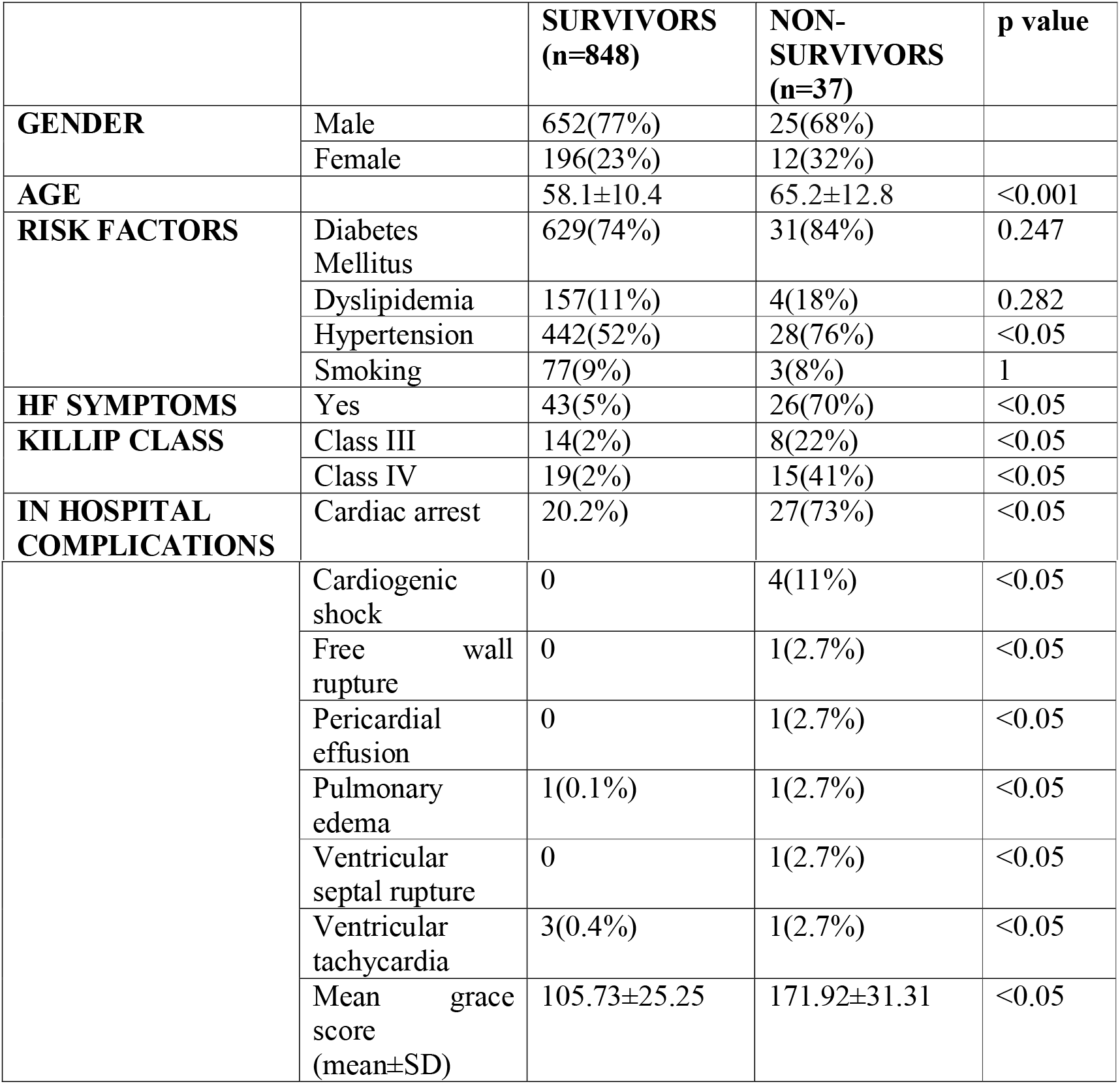
Outcome distribution of the demographic and clinical profiles of patients.

## DISCUSSION

The COVID-19 pandemic in 2019 affected approximately 20 million patients worldwide and 2.0 million cases from India. Lockdowns were employed to prevent the spread of the pandemic. However, it had an unintentional impact on access to healthcare, including acute cardiovascular care, especially acute coronary syndrome (ACS). Observational studies have shown a decrease in hospital admissions for AMI in several developed countries during the pandemic period. While coronavirus disease 2019 (COVID-19) is known to present with respiratory symptoms, it can have multiple system complications, such as cardiovascular manifestations. ^(6)^ In our study, 58% of patients presented early presenters, and 42% presented late presenters. Among the late presenters, 6% did not survive, whereas in early presenters, this number was reduced to 3%.

Our study coincides with a previous study which revealed that COVID-19 has high rates of fatality in older individuals, which might be due to the loss of endothelial function and endogenous cardioprotective mechanisms, resulting in an increased prevalence of cardiovascular disorders. (7) Similarly, in our study, the mean age of the study population was 58.45 ± 10.61 years, ranging from 27--85 years. Additionally, our study revealed a significant difference in the mean age of nonsurvivors compared with survivors.

A study conducted in 2019 revealed that the incidence of COVID-19 was greater in people who were predisposed to comorbidities such as diabetes mellitus and hypertension, as these people presented reduced ACE2 expression and upregulated angiotensin II proinflammatory signaling, which caused ACE-i/ARB treatment to be corrective; however, in patients with COVID-19, SARS-CoV-2 binds to ACE2, greatly exaggerating the proinflammatory background. ^(8)^ Alternatively, the release of circulating cytokines during severe systemic inflammatory stress can lead to atherosclerotic plaque instability and rupture^(9).^ Our study aligns with this study, in which 660 (75%) patients had diabetes mellitus and 470 (53%) of the population were hypertensive.

Another study revealed that SARS-CoV-2 infection may lead to the development of HF through direct acute cardiac injury or through the progression of cytokine storms, which depresses cardiomyocyte function and cardiac contractility. In our study, 70% of the people who did not survive showed heart failure symptoms, whereas 5% of the people who survived did. Anti-heart failure drugs, mainly digoxin and carvedilol, may attenuate the development of HF by reducing the infectivity of SARS-CoV-2 and preventing the development of cytokine storms in severely affected COVID-19 patients. ^(10)^

Inflammation in the vascular system can result in diffuse microangiopathy with thrombosis. Inflammation in the myocardium can result in myocarditis, heart failure, cardiac arrhythmias, acute coronary syndrome, rapid deterioration, and sudden death. Similarly, we analyzed in-hospital complications and their outcomes during the hospital stay. Among the 44 patients who had complications, 29 (4%) had cardiac arrest, and 4 had ventricular tachycardia and cardiogenic shock. We also noted in-hospital deaths in 37 (4.18%) patients. ^(11)^

Various observational studies have shown a decrease in hospital admissions for AMI in several developed countries during the pandemic period. Another study^(12,13)^ was conducted to determine the impact of increased mortality and worse cardiac outcomes of AMI during the early COVID-19 pandemic. They reported that mortality was greater during the early COVID-19 phase than during the pre-COVID-19 phase. They reported that the time from symptom onset to first medical contact was prolonged in all AMI patients during early COVID-19. Major cardiac complications after AMI occur significantly more often, and cardiac recovery is worse in patients with early-stage COVID-19. ^(12)^

During the first phase of the COVID-19 pandemic, there was a marked decline in acute cardiovascular hospitalizations, and patients who were admitted had shorter lengths of stay. These data substantiate concerns that acute care for cardiovascular conditions may be delayed, deferred, or abbreviated during the COVID-19 pandemic^(8)^. We propose that a delay between symptom onset and initial medical consultation resulted primarily from public health concerns related to the pandemic. Furthermore, patients who sought care earlier were more likely to exhibit severe symptoms, which prompted prompt medical attention. This pattern is also evident in the observed differences in patient survival rates and the incidence of in-hospital complications.

### Clinical implications

This study highlights the critical impact of delayed hospital presentation among STEMI patients during the COVID-19 pandemic, demonstrating a clear association between late-presentation, multivessel coronary disease, and increased in-hospital mortality. This finding emphasizes the importance of maintaining uninterrupted access to emergency cardiac care during public health crises and reinforces the need for early recognition, timely intervention, and patient education to improve cardiovascular outcomes in similar future scenarios.

### Strengths of the study

- This study included a large cohort of 885 patients diagnosed with ST-segment elevation myocardial infarction (STEMI) during the COVID-19 pandemic, providing robust statistical power for evaluating clinical patterns and outcomes.
- Data were collected from a tertiary care center with standardized diagnostic and management protocols, ensuring consistency in patient assessment, ECG interpretation, and coronary angiography findings.
- The study comprehensively analysed demographic factors, risk profiles, ECG characteristics, angiographic patterns, and in-hospital outcomes, allowing for a multidimensional understanding of STEMI presentations during the pandemic period.
- The inclusion of early versus late presenters offers valuable insights into how delayed presentation impacted complications, vessel involvement, and mortality, which is particularly relevant in pandemic-related healthcare disruptions.

### Limitations

- The study was retrospective and single center, which may limit the generalizability of the findings to other populations and healthcare settings.
- Selection bias cannot be excluded, as only hospitalized patients were included; those who died before reaching the hospital or were treated elsewhere were not captured.
- The study did not evaluate long-term outcomes or post-discharge mortality, focusing only on in-hospital events.
- Because of the pandemic context, variations in healthcare accessibility and treatment delays may have influenced outcomes independently of patient or disease characteristics.

## CONCLUSION

COVID-19 is an eye opener for healthcare workers to develop a necessary disease algorithm such that patients with symptoms of disease can reach healthcare facilities in time to prevent complications. Based on the results of our study, patients with more severe symptoms managed to reach the hospital early despite the pandemic. This was due to the severity of their symptoms resulting in their overall outcome. In addition, late presenters had milder symptoms, resulting in delayed contact with healthcare facilities and a greater chance of developing morbidity and mortality.

In addition, 5% of the late presenters developed cardiac arrest as well as other complications, such as ventricular free wall rupture and pericardial effusion, which was not noted in early presenters.

We also noted that late presenters mostly had double-or triple-vessel diseases, while most of the early presenters had single-vessel disease, and the likelihood of in-hospital complications and mortality was greater in late presenters.

## Data Availability

Data related to this study is available at the Open Science Framework

https://osf.io/zcahy/overview?view_only=15d6c521b5f947a8a6ec5981042a08d9

## DECLARATIONS

## 1. Abbreviations

ACS: acute coronary syndrome
AMI: acute myocardial infarction
ARDS: acute respiratory distress syndrome
AWMI: Anterior wall myocardial infarction
BMI: Body mass index
BSA: Body surface area
CHB: Complete Heart Block
COVID-19: Coronavirus Disease 2019
DVD: Double vessel disease
ECG: Electrocardiogram/Electrocardiographic
HF: Heart failure
IWMI: Inferior wall myocardial infarction
LBBB: Left Bundle Branch Block
MI: Myocardial infarction
SARS-CoV-2: Severe acute respiratory syndrome coronavirus-2
SD: standard deviation
STEMI: ST-Segment Elevation Myocardial infarction
SVD: single vessel disease
TIMI: Thrombolysis in Myocardial Infarction
TVD: Triple Vessel Disease

## 2. Human Ethics and Consent to Participate

This study adhered to the Declaration of Helsinki and was conducted after the approval of the Institutional Ethics Committee (IEC) of Kasturba Medical College, Mangalore (IEC KMC MLR 07-2021/242). This was record based retrospective study. Hence informed consent was not considered. This study caused no harm to the participants, researchers or the health system. This article reported following the Strengthening the Reporting of Observational Studies in Epidemiology (STROBE) guidelines for Observational studies.

## 3. Consent for publication

Not Applicable

## 4. Availability of data

Data related to this study is available at the Open Science Framework from https://osf.io/zcahy/overview?view_only=15d6c521b5f947a8a6ec5981042a08d9

## 7. Author’s contribution

D.N: Conceptualization, data collection, methodology, formal analysis, writing original draft H.H : Conceptualization, supervision, visualization & validation P.K: Conceptualization, supervision, review & editing M.C: Review & editing A.P: Review & editing C.N: Methodology, Editing R.D: Methodology, Editing

All authors have approved the submitted version

## 8. Acknowledgments

Not Applicable

## 9. Clinical Trial Number

Not applicable

